# Scalable system-wide CYP2C19 pharmacogenomic testing reveals 38% excess incidence of adverse events in metabolizers receiving inappropriate prescriptions

**DOI:** 10.1101/2025.03.13.25323700

**Authors:** Natalie Telis, Douglas Stoller, Christopher N. Chapman, C. Anwar A. Chahal, Daniel P. Judge, Douglas A. Olson, Joseph J. Grzymski, Teresa Kruisselbrink, Nicole L. Washington, Elizabeth T. Cirulli

## Abstract

**Objective:** In spite of evidence and recommendations reflecting the importance of pharmacogenomic testing, most prescriptions are still given without testing. We demonstrate the real world implications of the use of testing and evaluate adverse events and outcomes in individuals who did not receive pharmacogenomic testing for clopidogrel.

**Methods:** We analyzed ~100K individuals with paired EHR and exome sequencing data from population health studies administered at multiple medical centers using the Helix Exome+^®^ assay. We inferred clopidogrel dosage by processing the prescription with an LLM. We identified all instances of individuals with at least one prescription that is not in concordance with their CYP2C19 genotype. Lastly, we identify instances of thrombosis using a comprehensive codeset based on ICD9, ICD10, and SNOMED terms.

**Results:** We identified 16,140 prescriptions of clopidogrel given to 3,853 participants. We found that 29% of these individuals have a mismatch between the recommended clopidogrel dosage guideline based on their CYP2C19 genotype and their actual prescribed daily dosage. 25% of poor metabolizers experienced thrombosis, with 40% occurring within 2 months of treatment. Poor and intermediate metabolizers receiving clopidogrel are much more likely to experience thrombosis and myocardial infarction (binomial *p-*value = 0.001).

**Conclusions:** We estimate a 38% excess of adverse events occur in poor and intermediate metabolizers relative to normal and rapid metabolizers. The lack of testing may be responsible for 1 thrombosis event per every ~30 people prescribed clopidogrel.

---

In spite of well-established clinical utility, best practices, and recommendations to use pharmacogenetic testing from the FDA (1), the American Heart Association (2) and the Clinical Pharmacogenomic Implementation Consortium (3), few people receive pharmacogenetic testing during clopidogrel treatment. Clopidogrel is an antiplatelet drug prescribed to prevent thrombosis, but *CYP2C19* is required to metabolize it into its active state. Inadequate *CYP2C19* function is therefore tied to a lack of protection against stroke, myocardial infarction (MI), and death in individuals taking clopidogrel.

We measure the real world consequences of the lack of pharmacogenetic testing through a retrospective study of patients prescribed clopidogrel. We study these adverse events and outcomes in individuals who did not receive pharmacogenetic testing, focusing on individuals whose treatment course is discordant with best practices for pharmacogenetic implementation. We demonstrate that inappropriate prescriptions are common and result in 22 extra thrombosis events and 16 extra MIs per 1000 individuals taking clopidogrel.

We study the Helix Research Network, an all-comers cohort from multiple medical sites that provides electronic health records alongside DNA sequencing using the Helix Exome+^®^ assay. This assay enables pharmacogenomic testing for this study as well as reanalysis to derive additional genomic information. We use the Exome+^®^ data to derive *CYP2C19* star alleles (4). We then use electronic medical records to identify individuals with at least one prescription of clopidogrel or the recommended alternative drugs prasugrel and ticagrelor. In these individuals, we identified thrombosis, MI and major adverse cardiac events (MACE) over three timepoints within a year of the first drug prescription, using a comprehensive codeset based on ICD9, ICD10, and SNOMED terms. Next, we identified dosage by processing the freetext prescription “sig” field with an LLM, which showed 99% accuracy against human-derived gold standard doses. We use these data to identify all instances of individuals with at least one prescription that is not in accordance with the CPIC clopidogrel use guidelines based on their *CYP2C19* genotype (3). All poor metabolizers, and all intermediate metabolizers taking a 75mg or lower dose of clopidogrel, were categorized as being inappropriately prescribed.

In our cohort of 101,883 individuals, we identify 16,140 prescriptions of either clopidogrel, prasugrel, or ticagrelor given to 4,057 distinct individuals. Of these, 29% are inappropriately prescribed clopidogrel. In particular, 26% have intermediate metabolic (IM) activity of CYP2C19, 97% of whom are taking an unadjusted dose of clopidogrel. Another 3% have poor metabolic (PM) CYP2C19 activity, and should not take clopidogrel at any dose. *CYP2C19* genotype proportions were identical between people taking clopidogrel and people taking alternative drugs, implying genotype is not related to which drug is taken in the present dataset (X^2^ = 3.6, *p* = 0.71). In inappropriately prescribed individuals, we observe an excess of thromboses, myocardial infarctions (MI), and a five point measure of major adverse cardiac events (MACE) including MI, stroke, cardiovascular mortality, unstable angina, and heart failure. A majority of adverse events occurred within 2 months of prescription start (see Fig1A, B, and C respectively). These trends were consistent within genetic similarity groups, were not explained by age or sex, were consistent in people with and without percutaneous coronary intervention (PCIs), and were confirmed with independent data on excess hospitalizations (data not shown).

**Figure 1.**
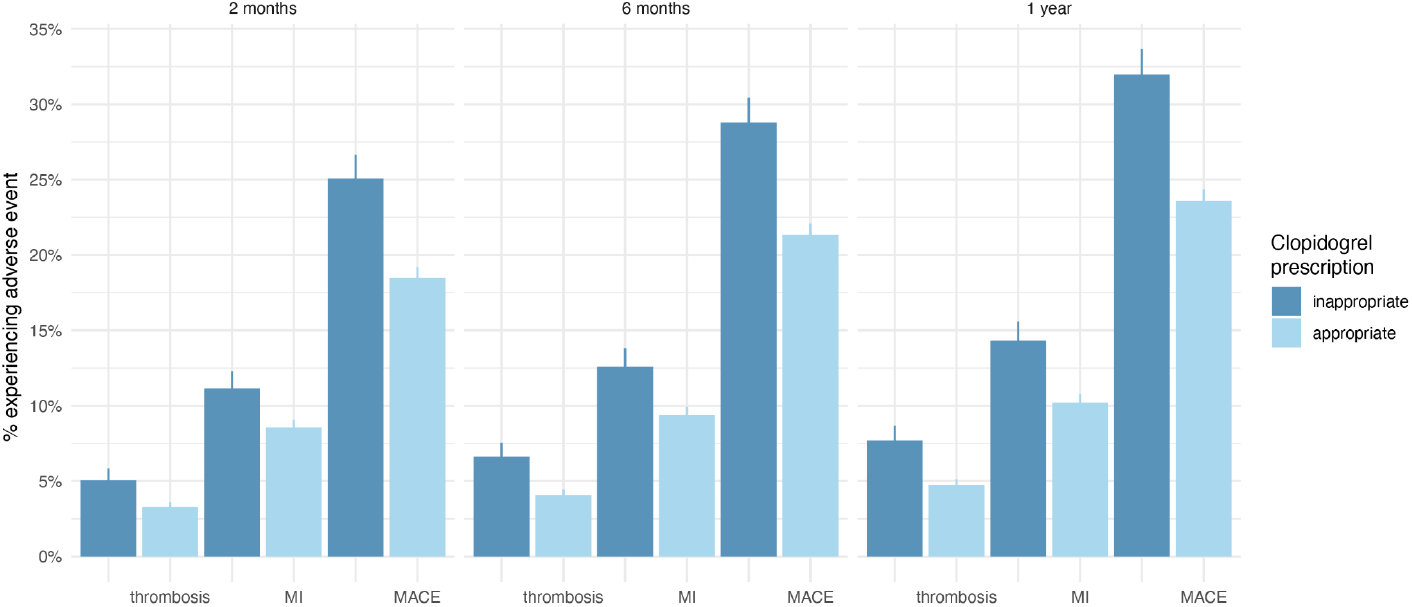
The incidence of cardiovascular morbidity in the Helix Research Network, occurring either 2 months (left), 6 months (middle), or within 1 year (right) of the first documented clopidogrel prescription. Thrombosis, myocardial infarction (MI) and major adverse cardiac events (MACE) are defined using a combination of ICD9, ICD10, ICD10CM and SNOMED terms. Individuals with events prior to prescription are not excluded, as the expectation is that a prior event may prompt intervention and clopidogrel prescription.

The utility of a PGx test relies on the genotypic information discriminating between multiple appropriate choices, such as alternate antiplatelet drugs. While we identified 423 individuals who received alternative antiplatelet drugs, most took them after taking clopidogrel and experiencing cardiac events: only 4 poor metabolizers took an alternate drug first, which did not provide sufficient power to distinguish outcome incidence.

## Discussion

In this study of an all-comers cohort, we demonstrate that the lack of pharmagenomically informed prescription is enormously impactful for patients taking clopidogrel. We find there is an excess incidence of severe adverse events in individuals who should not take clopidogrel, causing a direct harm to patients that could be prevented by genetic testing. We estimate that per 1000 people tested, there is a cost of $700k attributable to these excess thromboses and MI (5). This quantifies the clinical utility, patient benefit, and monetary value of pharmacogenetic testing in a real patient population receiving the current standard of care.

To create this cohort, we demonstrate the ability to call pharmacogenetically relevant genotypes in tens of thousands of patients with Exome+^®^ sequencing, enabling diversely applicable pharmacogenetic testing at scale. Moreover, the proof of principle with Exome+^®^ data enables clinically applicable genetic testing far beyond *CYP2C19*, because once the genetic data are stored, they can be referred back to for many future applications. Our study addresses the tangible benefit of genomic testing when delivering pharmacogenetic testing to a healthcare system at scale. This affirms the feasibility and impact of an accessible genome as part of the standard of clinical care.

## Data Availability

The Helix cohorts data and code associated with these analyses are available to qualified researchers upon request and with permission of the participating health systems. The Helix cohorts encourage and collaborate with scientific researchers on an individual basis. Examples of restrictions that will be considered in requests to data access include but are not limited to (1) whether the request comes from an academic institution in good standing and will collaborate with our team to protect the privacy of the participants and the security of the data requested, (2) type and amount of data requested, (3) feasibility of the research suggested, and (4) amount of resource allocation required to support the collaboration. Any correspondence and data availability requests related to this data should be addressed to N.T.

